# PHYSICIANS’ ADHERENCE TO GUIDELINES ON MEDICAL THERAPY FOR HEART FAILURE AND PATIENTS’ OUTCOMES IN A KENYAN REFERRAL HOSPITAL

**DOI:** 10.1101/2023.10.31.23297731

**Authors:** Willy Mucyo, Jasmit Shah, Jeilan Mohamed, Mohamed Hasham Varwani, Mzee Ngunga

**Affiliations:** Department of Medicine, Aga Khan University, Nairobi, Kenya; Brain and Mind Institute, Aga Khan University, Nairobi, Kenya

**Keywords:** Heart failure with reduced ejection fraction, guideline-directed medical therapy, guideline adherence, Sub-Saharan Africa

## Abstract

**Background:** Adherence to guidelines on prescription and uptitration of GDMT for HFrEF is associated with reduced mortality and hospitalization. Published data on physicians’ GDMT prescription in sub-Saharan Africa is scarce. In addition, there is a need for data on patients’ characteristics, treatment, and outcomes in this region.

**Objective:** To determine physicians’ level of adherence to guidelines on prescription and uptitration of medical therapy for HFrEF at AKUHN, a referral hospital in Nairobi, Kenya.

**Methods:** We reviewed 280 files of all HFrEF patients admitted over a 3-year period. Detailed patients’ characteristics and outcomes were analyzed. We calculated the Guideline Adherence Index (GAI) and the QUality of Adherence to guideline recommendations for LIFe-saving treatment in heart failure (QUALIFY) scores. From worst to best, GAI ranges from 0 to 100%, while QUALIFY scores were categorized as poor, moderate, or good.

**Results:** The median age (IQR) was 63 years (53,74); 165(58.9%) were male; and 207(74.2%) were black; 98(35%) had ischemic heart disease; 153(54.6%) had hypertension, and 101(36.1%) had diabetes. At six months follow-up, 43.8% of patients had been readmitted at least once and 8.8% had died. GAI at discharge were 66.2%, 71.7%, and 42.6% for ACEI/ARNI/ARBs, B-blockers, and MRAs, respectively. At 6 months, the scores were 86.3%, 84.4%, and 61.2%, respectively. GAI for SGLT2is was 38.9% at 6 months. The proportions for good QUALIFY scores for ACEI/ARNI/ARBs, B-blockers, and MRAs were 35.8%, 38.5%, and 9.5%, respectively. Uptitration to ≥ 50% of target dose was done in 51.9%, 48.7%, and 7.9% of patients for ACEI/ARNI/ARBs, B-blockers, and MRAs, respectively.

**Conclusion:** Physicians’ level of adherence to prescription and up-titration of GDMT was satisfactory for ACEI/ARNI/ARBs and B-blockers, however, it was poor for SGLT2is and MRAs. There is a need for regular surveys on prescription and uptitration of GDMT with a special attention to MRAs and SGLT2is.

## INTRODUCTION

### Background

It is estimated that 64.3 million people live with heart failure (HF) worldwide.[1] The prevalence of heart failure in adult population is estimated at 1% to 2% in developed countries, and up to 5% in low and middle-income communities in these countries. The evidence suggests that the number of HF patients in low and middle-income countries is rising.[2] A review of HF in Africa found the most common causes to be hypertensive heart disease (HHD), ischemic heart disease (IHD), and dilated cardiomyopathy (DCM). In sub-Saharan Africa (SSA), the most prevalent cause was HHD, in contrast with IHD in north Africa [3] A single-center study conducted in Kenya reported cardiomyopathy as the most common cause of HF, closely followed by HHD.[4] HF accounts for 9.4 – 15% of all hospital admissions in SSA and patients tend to be younger compared to those in developed countries.[5]

In 2018 the Kenyan ministry of health published guidelines for management of cardiovascular diseases. Key recommendations for heart failure with reduced ejection fraction (HFrEF) treatment (i.e., use of angiotensin receptor blockers [ACEIs] / angiotensin receptor-neprilysin inhibitor [ARNI] / angiotensin receptor blockers [ARBs], beta blockers [B-blockers], and mineralocorticoid receptor antagonists [MRAs]) do not differ from those by the American Heart Association/American College of Cardiology (AHA/ACC) and the European Society of Cardiology(ESC), except for the addition of sodium-glucose cotransporter-2 inhibitors (SGLT2is) in the 2021 ESC guidelines.[6]–[9] The guidelines strongly recommend (Class IA) that, provided that there are no contra- indications, medications in the three aforementioned classes should be started before discharge and gradually uptitrated (every 2 weeks for ACEI/ARNI/ARBs and B-blockers and 4-8 weeks for MRAs) to target doses or maximally tolerated doses.[10] The recommendations are based on a large body of evidence from randomized clinical trials supporting the use of guideline-directed medical therapy (GDMT) in HFrEF[11]–[23].

### Physicians’ guideline adherence

Although measuring guideline adherence is not a perfect science, efforts have been made to quantify it. One of the three domains of the ESC quality indicators for HF care solely focuses on guideline adherence. This quality indicator, also referred to as Guideline Adherence Index (GAI) is measured separately for each IA drug class. The score is expressed as a proportion of patients with prescriptions out of eligible patients (see supplement III).[10]Eligibility is defined as lack of contraindications or reasons for discontinuation.[10] Supplement I contains the list of contraindications and reasons for discontinuation, for each drug class, as published by the ESC. GAI is a simple score that does not require robust data analysis and can be used in various healthcare settings. However, the score is limited as it does not take into account dose uptitration. In addition, GAI cannot be used to assess variations between individual or patient subgroups.

National and international HF registries have used different methods to assess guideline adherence. The QUality of Adherence to guideline recommendations for LIFe-saving treatment in heart failure (QUALIFY) investigators calculated guideline adherence scores for each drug class for each patient. The scores varied from 0 (poor) to 1 (good). The scores were calculated as follows (see supplement III): lack of prescription for an eligible patient = 0 points; prescription of < 50% of the target dose for an eligible patient = 0.5 points; prescription of >= 50% of the target dose for an eligible patient = 1 point; prescription for an ineligible patient = 0 points; and lack of prescription for an ineligible patient = 1 point.[24], [25] The overall guideline adherence level for three drug classes was qualified as good if all indicated drugs were prescribed, moderate if > 50% of indicated medications were prescribed, and poor if ≤ 50% of indicated medications are prescribed. The QUALIFY score provides adherence scores for individual patients and may be used to assess subgroup differences. Furthermore, it takes into account dose uptitration.

The Korean Acute Heart Failure registry (KorAHF) used a modified version of the QUALIFY score which adds anticoagulation for stroke prevention in atrial fibrillation. Three levels of adherence were defined: good (prescription of all indicated medications), moderate (half or more of the indicated medications), and poor (prescription of less than half of indicated medications). This scoring system considers differences between patients but does not account for dosage uptitration. In a sense, it is more complex compared with GAI and simpler compared with the original QUALIFY.

Guideline adherence is associated with reduced HF mortality and HF hospitalization. The QUALIFY study which has a registry of 6118 ambulatory HFrEF patients in 549 centers in 36 countries, found that physicians’ guideline adherence was associated with reduced HF death and HF hospitalization or CV death.[25] A recent large systematic review and meta-analysis of 45866 patients from 11 multi-center studies, including the QUALIFY study, found significant association between good adherence and reduced all-cause mortality.[26] In KorAHF registry which has 5625 patients with HF and atrial fibrillation, guideline adherence at discharge was associated with significantly reduced mortality at 60-day and 1-year follow-up.[27] The evidence shows that guideline adherence at discharge is as important as long-term adherence.[25], [27] Uptitration to more than 50% of the target dose is also essential as it has been shown to decrease mortality in HF patients.[26]

The QUALIFY registry includes 19 and 15 centers in two African countries, Egypt and Morocco respectively. This survey found satisfactory and comparable guideline adherence scores in all geographic regions except for central and eastern Europe which had significantly lower scores. In the north African region, the guideline adherence was good, moderate, and poor in 71%, 22%, and 7% of patients, respectively, and the global adherence was 67%, 25%, and 8%, respectively.[24] The recent large systematic review and meta-analysis on physicians’ adherence to HF guidelines reported a 80.9% prescription rate for ACEI/ARBs, 78.0% for B-blockers, and 47.4% for MRAs.[26] Two studies conducted in Nigeria and Ethiopia found prescription rates of 83% and 79.8%, 48% and 79.4%, and 41% and 49%, for ACEI/ARBs, B-blockers, and MRAs, respectively.[28], [29]

### Justification of the study

Since physicians’ guideline adherence has been shown to reduce HFrEF mortality and hospitalization, societal guidelines recommend assessing the adherence as a major quality indicator of HFrEF care. The present study determines the current guideline adherence at AKUHN which may help improve HFrEF care. Furthermore, it contributes to the currently scarce published data on HFrEF patients’ characteristics, treatment, and outcomes in the SSA region.

### Study objectives

The primary objective is to determine physicians’ level of adherence to guidelines on prescription and uptitration of medical therapy for HFrEF at AKUHN. Secondary objectives are to determine the association between adherence scores and mortality rate at 6 months and to determine the association between adherence scores and readmission for heart failure within 6 months.

## METHODS

### Study population

The study was conducted at the Aga Khan University Hospital, Nairobi, a tertiary care, teaching and referral facility. It included adult patients (18 years and above) with documented diagnosis of heart failure with reduced LVEF (≤40%). We excluded patients who died during the first hospitalization and those with missing LVEF data.

As shown in *figure 1*, between January 2020 and December 2022, we identified and reviewed 736 admissions files for “congestive heart failure (I50.0), left ventricular failure (I50.1), and heart failure unspecified (I50.9)”. Of these files, the following were excluded: 227 were not heart failure conditions, 86 were heart failure with preserved ejection fraction, 30 were heart failure with mid-range ejection fraction, and 38 had no ejection fraction (EF) record. Of the remaining 355 HFrEF files, 75 were readmissions, hence 280 unique HFrEF patients.

**Figure 1:**
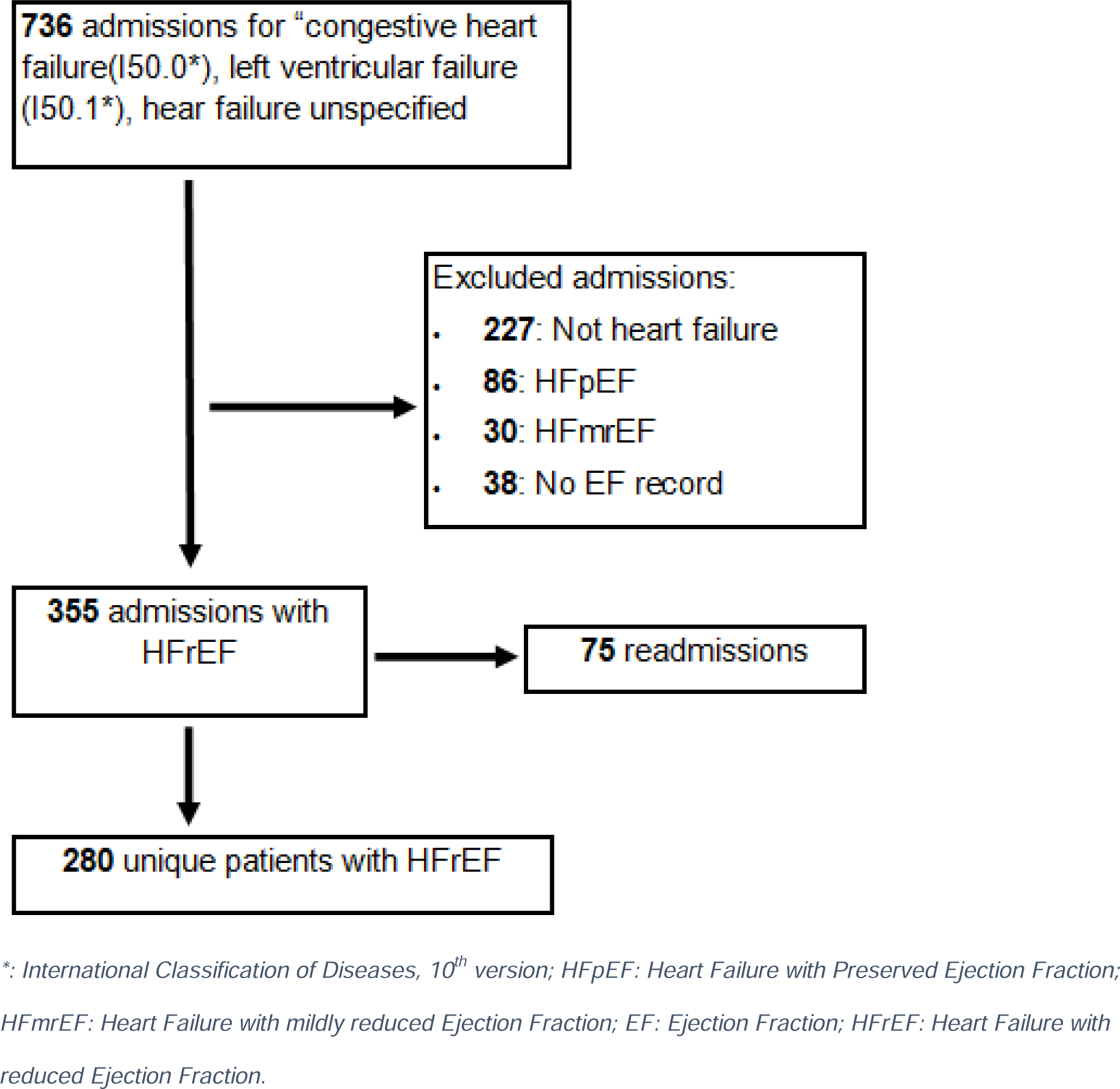
Patients selection.

### Data analysis

Continuous data was presented as medians with interquartile ranges or means with standard deviations, whereas categorical data was presented as frequencies with percentages. The levels of guideline adherence were calculated using GAI and QUALIFY scores as published by the ESC and QUALIFY investigators, respectively (supplement III). GAI were reported as percentages and individual QUALIFY scores were reported as follows: poor (score = 0), moderate (score = 0.5), and good (score = 1) for each drug class for each patient. The overall guideline adherence was qualified as good if all indicated drugs were prescribed, moderate if > 50% of indicated medications were prescribed, and poor if ≤ 50% of indicated medications are prescribed. Regarding exploratory outcomes, univariate analysis was done using univariate logistic regression for categorical or continuous data. Furthermore, multivariate logistic regression analysis was used in identifying independent associations between independent variables.

### Ethical consideration

Ethical clearance was obtained from the Aga Khan University Nairobi Institutional Scientific Ethics Review Committee.

## RESULTS

### Patients’ characteristics

As shown in *Table 1*, the median age (interquartile range) was 63 years (53, 74); 165 (58.9%) patients were male; and 207 (74.2%) were black. Most patients (68.6%) were admitted in non-critical units. The most prevalent primary cause of HF was IHD, accounting for 35% of cases, followed by HHD (21.1%). 182 (65%) patients were under general practice, as defined by the hospital, while 93 (35%) were under private practice. 153 patients (54.6%) had hypertension, 101 (36.1%) had diabetes, and 48 (17.2%) had atrial fibrillation. 131 (46.8%) patients had undergone coronary angiogram at some point in the past and 53 (19%) had a history of myocardial infarction. 86.3% of patients had functional limitation classified as NYHA class II or III. The median LVEF was 30% (10, 35) and the prevalence of atrial fibrillation on ECG was 16.4%. The initiation rate of cardiac rehabilitation initiation before discharge was 10.5%.

**Table 1:**
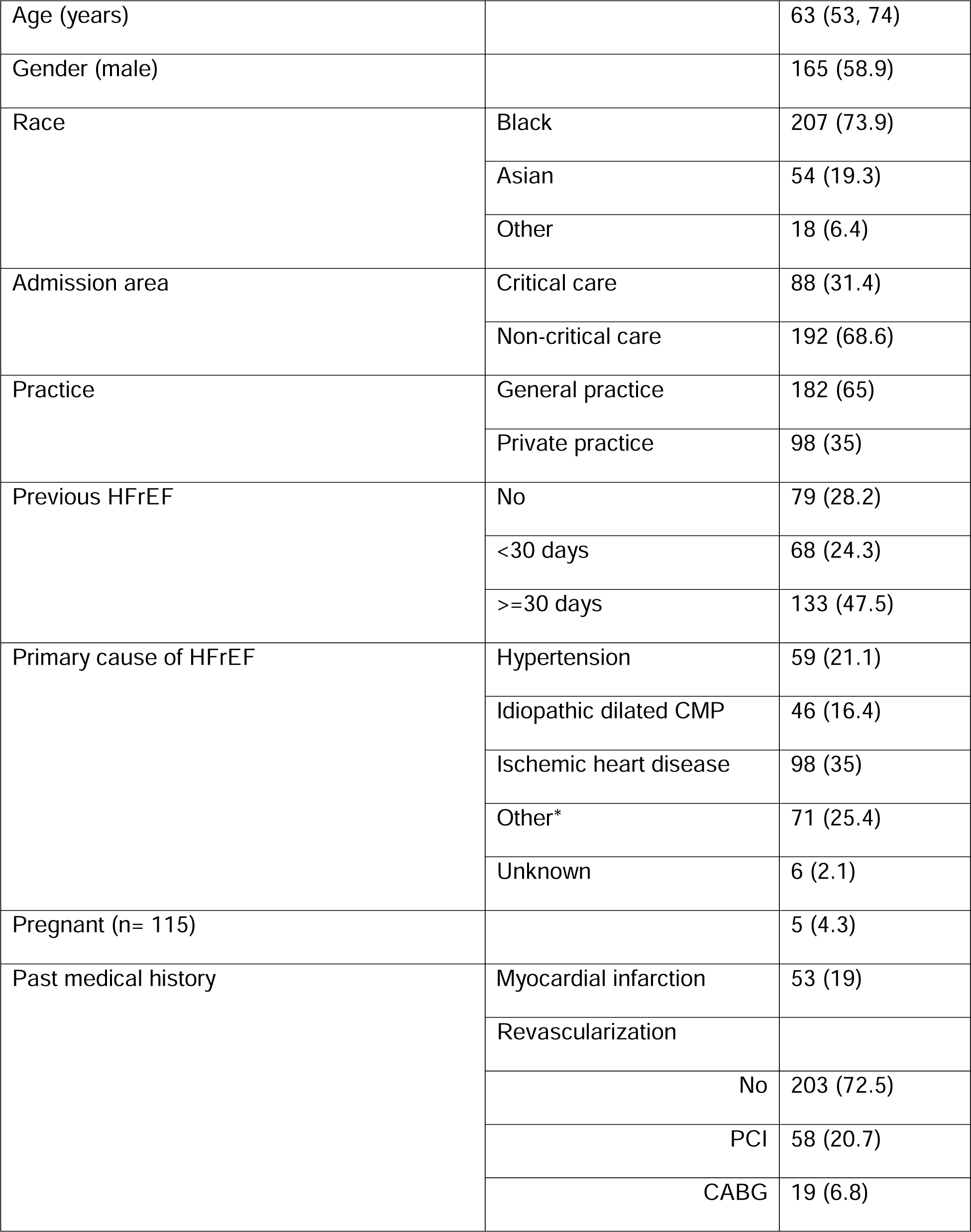
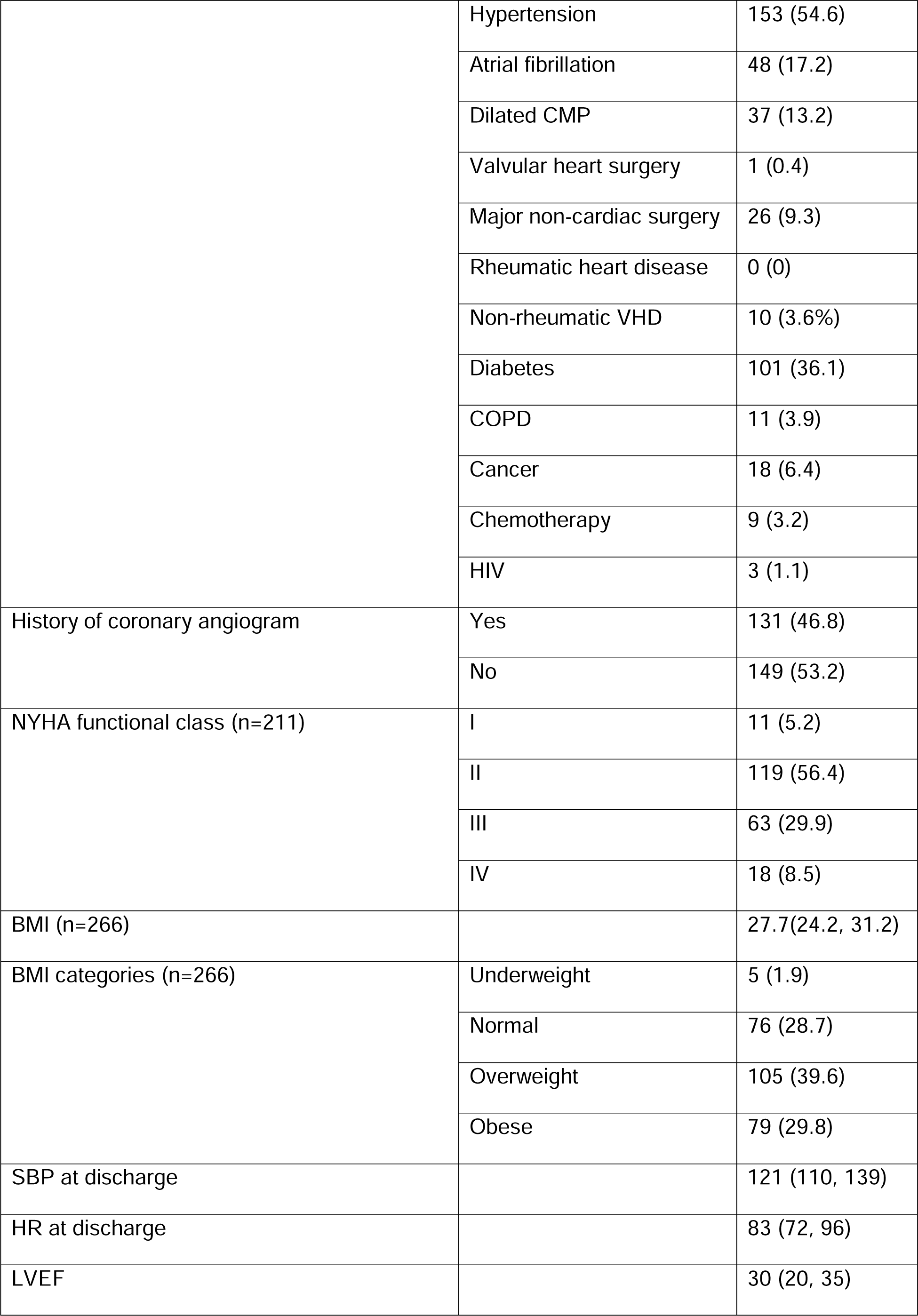
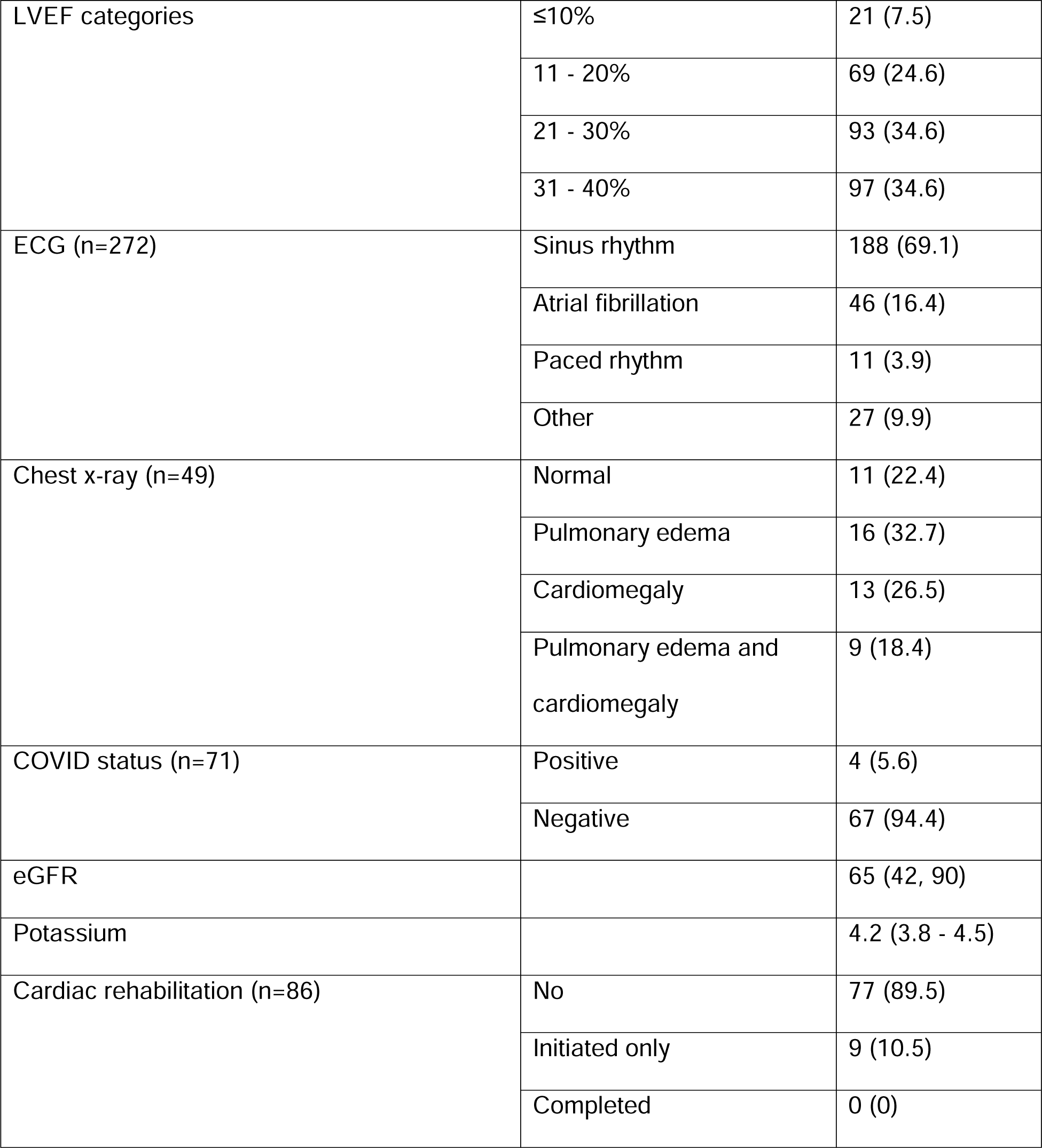
Patients’ characteristics.

|  |  |  |
| --- | --- | --- |
| Age (years) |  | 63 (53, 74) |
| Gender (male) |  | 165 (58.9) |
| Race | Black | 207 (73.9) |
|  | Asian | 54 (19.3) |
|  | Other | 18 (6.4) |
| Admission area | Critical care | 88 (31.4) |
|  | Non-critical care | 192 (68.6) |
| Practice | General practice | 182 (65) |
|  | Private practice | 98 (35) |
| Previous HFrEF | No | 79 (28.2) |
|  | <30 days | 68 (24.3) |
|  | >=30 days | 133 (47.5) |
| Primary cause of HFrEF | Hypertension | 59 (21.1) |
|  | Idiopathic dilated CMP | 46 (16.4) |
|  | Ischemic heart disease | 98 (35) |
|  | Other* | 71 (25.4) |
|  | Unknown | 6 (2.1) |
| Pregnant (n= 115) |  | 5 (4.3) |
| Past medical history | Myocardial infarction | 53 (19) |
|  | Revascularization |  |
|  | No | 203 (72.5) |
|  | PCI | 58 (20.7) |
|  | CABG | 19 (6.8) |
|  | Hypertension | 153 (54.6) |
|  | Atrial fibrillation | 48 (17.2) |
|  | Dilated CMP | 37 (13.2) |
|  | Valvular heart surgery | 1 (0.4) |
|  | Major non-cardiac surgery | 26 (9.3) |
|  | Rheumatic heart disease | 0 (0) |
|  | Non-rheumatic VHD | 10 (3.6%) |
|  | Diabetes | 101 (36.1) |
|  | COPD | 11 (3.9) |
|  | Cancer | 18 (6.4) |
|  | Chemotherapy | 9 (3.2) |
|  | HIV | 3 (1.1) |
| History of coronary angiogram | Yes | 131 (46.8) |
|  | No | 149 (53.2) |
| NYHA functional class (n=211) | I | 11 (5.2) |
|  | II | 119 (56.4) |
|  | III | 63 (29.9) |
|  | IV | 18 (8.5) |
| BMI (n=266) |  | 27.7(24.2, 31.2) |
| BMI categories (n=266) | Underweight | 5 (1.9) |
|  | Normal | 76 (28.7) |
|  | Overweight | 105 (39.6) |
|  | Obese | 79 (29.8) |
| SBP at discharge |  | 121 (110, 139) |
| HR at discharge |  | 83 (72, 96) |
| LVEF |  | 30 (20, 35) |
| LVEF categories | ≤10% | 21 (7.5) |
|  | 11 - 20% | 69 (24.6) |
|  | 21 - 30% | 93 (34.6) |
|  | 31 - 40% | 97 (34.6) |
| ECG (n=272) | Sinus rhythm | 188 (69.1) |
|  | Atrial fibrillation | 46 (16.4) |
|  | Paced rhythm | 11 (3.9) |
|  | Other | 27 (9.9) |
| Chest x-ray (n=49) | Normal | 11 (22.4) |
|  | Pulmonary edema | 16 (32.7) |
|  | Cardiomegaly | 13 (26.5) |
|  | Pulmonary edema and cardiomegaly | 9 (18.4) |
| COVID status (n=71) | Positive | 4 (5.6) |
|  | Negative | 67 (94.4) |
| eGFR |  | 65 (42, 90) |
| Potassium |  | 4.2 (3.8 - 4.5) |
| Cardiac rehabilitation (n=86) | No | 77 (89.5) |
|  | Initiated only | 9 (10.5) |
|  | Completed | 0 (0) |

*Table 2* shows patients’ characteristics at 6 months follow-up. Most patients (61.1%) had a heart rate equal to or above 70 bpm. Most patients (89.7%) were classified as NYHA functional class I or II. Within the 6 months of follow-up, 63 (36.1%) patients were readmitted once, 10 (6.9%) were readmitted twice, and 1 (0.7%) was readmitted three times. 16 (8.8%) patients died during the 6-month follow-up period. Of those who died, 7 (43.8%) died of non-cardiovascular cause, 6 (37.5%) died of cardiovascular cause, and 3 (18.8%) died of unknown cause.

**Table 2:**
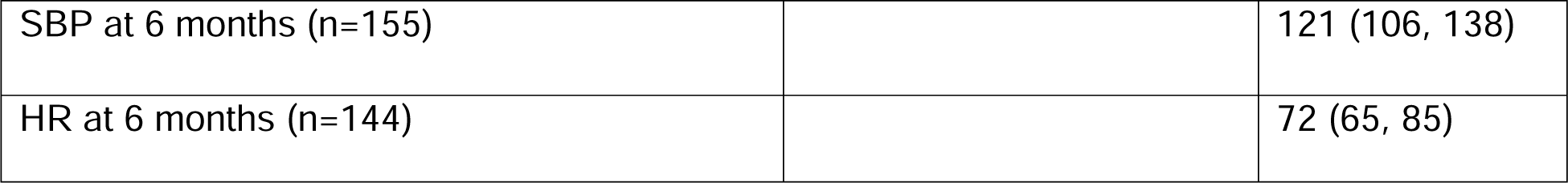
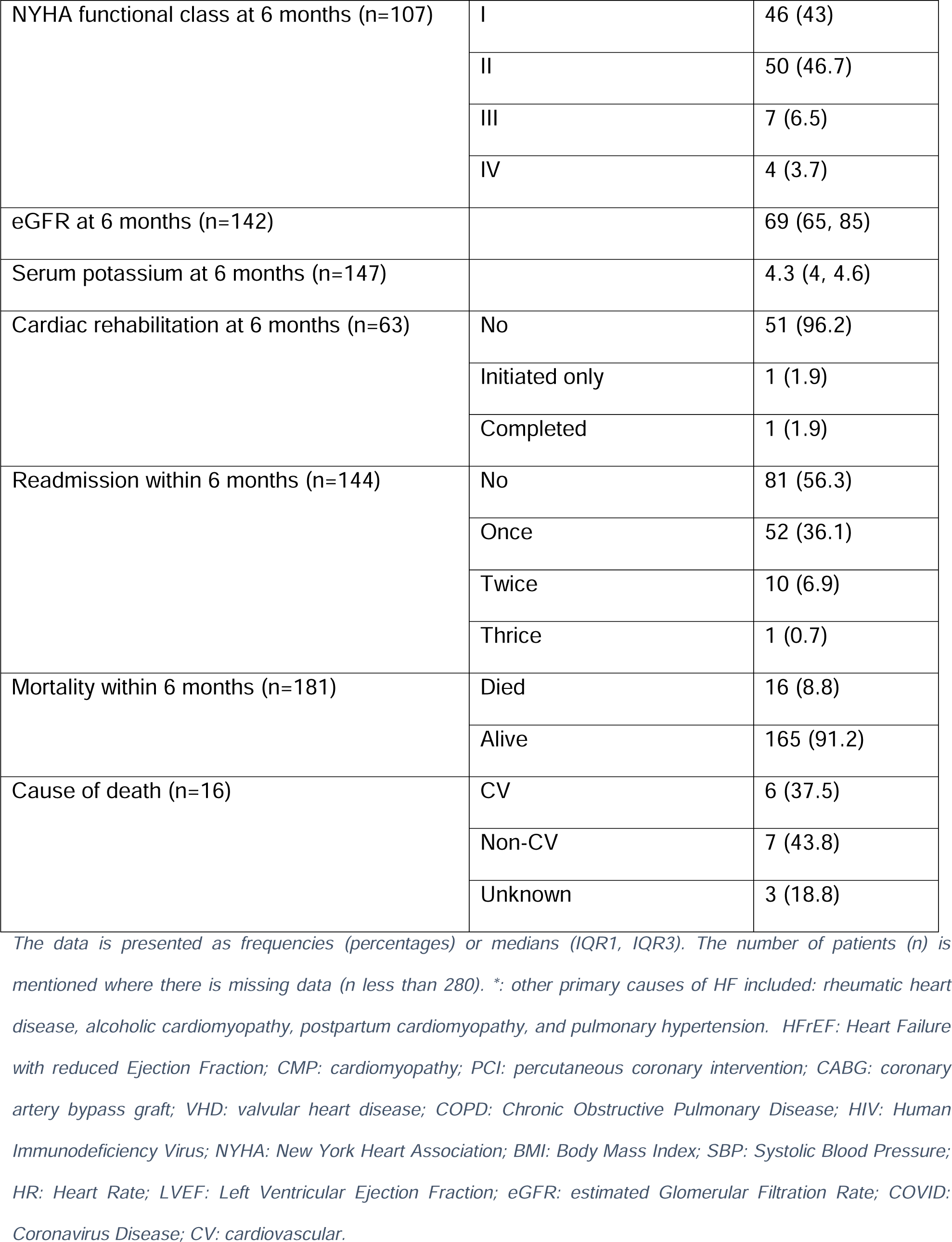
Six months follow-up.

|  |  |  |
| --- | --- | --- |
| SBP at 6 months (n=155) |  | 121 (106, 138) |
| HR at 6 months (n=144) |  | 72 (65, 85) |
| NYHA functional class at 6 months (n=107) | I | 46 (43) |
|  | II | 50 (46.7) |
|  | III | 7 (6.5) |
|  | IV | 4 (3.7) |
| eGFR at 6 months (n=142) |  | 69 (65, 85) |
| Serum potassium at 6 months (n=147) |  | 4.3 (4, 4.6) |
| Cardiac rehabilitation at 6 months (n=63) | No | 51 (96.2) |
|  | Initiated only | 1 (1.9) |
|  | Completed | 1 (1.9) |
| Readmission within 6 months (n=144) | No | 81 (56.3) |
|  | Once | 52 (36.1) |
|  | Twice | 10 (6.9) |
|  | Thrice | 1 (0.7) |
| Mortality within 6 months (n=181) | Died | 16 (8.8) |
|  | Alive | 165 (91.2) |
| Cause of death (n=16) | CV | 6 (37.5) |
|  | Non-CV | 7 (43.8) |
|  | Unknown | 3 (18.8) |

*The data is presented as frequencies (percentages) or medians (IQR1, IQR3). The number of patients (n) is mentioned where there is missing data (n less than 280). \*: other primary causes of HF included: rheumatic heart disease, alcoholic cardiomyopathy, postpartum cardiomyopathy, and pulmonary hypertension. HFrEF: Heart Failure with reduced Ejection Fraction; CMP: cardiomyopathy; PCI: percutaneous coronary intervention; CABG: coronary artery bypass graft; VHD: valvular heart disease; COPD: Chronic Obstructive Pulmonary Disease; HIV: Human Immunodeficiency Virus; NYHA: New York Heart Association; BMI: Body Mass Index; SBP: Systolic Blood Pressure; HR: Heart Rate; LVEF: Left Ventricular Ejection Fraction; eGFR: estimated Glomerular Filtration Rate; COVID: Coronavirus Disease; CV: cardiovascular.*

### GAI and QUALIFY scores

As shown in *Table 3*, at discharge, the highest prescription rate for eligible patients was with B-blockers while the lowest was with MRAs. Out of 272 patients who were eligible for B-blockers, 195 had prescriptions, resulting in a GAI of 71.8%. The GAI for ACEI/ARNI/ARBs was 66.2% (out of 234 patients who were eligible, 155 had prescriptions). As for MRAs, only 112 of 263 patients who were eligible had prescriptions (GAI: 42.6%).

**Table 3:**
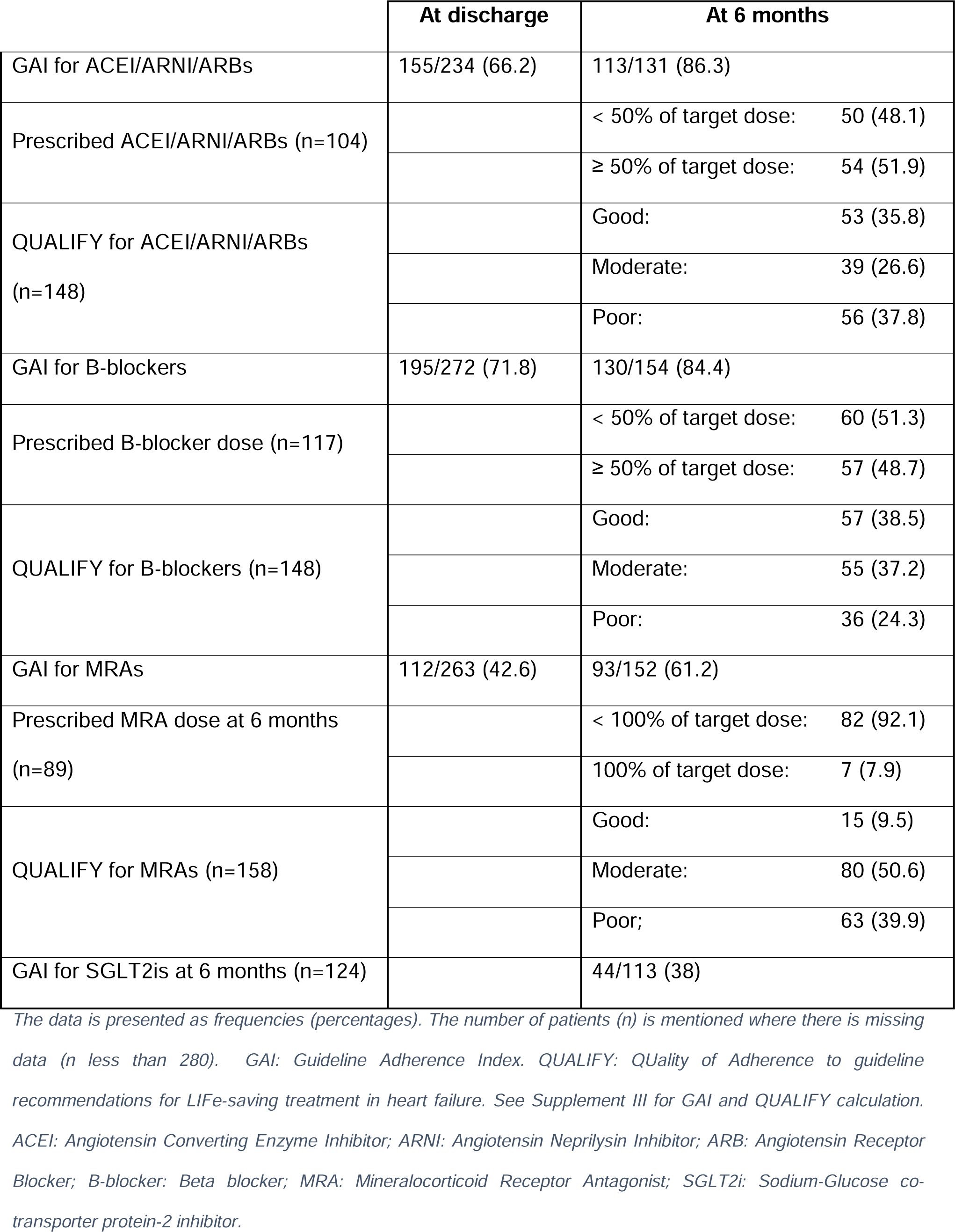
GAI and QUALIFY scores at discharge and at 6 months.

GAI and QUALIFY scores were calculated at 6 months follow-up. The highest GAI was with ACEI/ARNI/ARBs at 86.3% (113 had prescriptions out of 131 who were eligible); and of those with prescriptions, 51.9% had ≥50% of the target dose. QUALIFY scores for ACEI/ARNI/ARBs were good, moderate, and poor in 35.8%, 26.6%, and 37.8% of patients, respectively. GAI for B-blockers was 84.4% (154 were eligible and 130 had prescriptions); among those with prescriptions, 48.7% had ≥50% of the target dose. QUALIFY scores for B-blockers were good, moderate, and poor in 38.5%, 37.2%, and 24.3% of patients, respectively, while they were 9.5%, 50.6%, and 39.9% for MRAs, respectively. GAI for MRAs was 61.2% (152 were eligible and 93 had prescriptions); and of those with prescriptions, only 7.9% had 100% of the target dose.

**Figure 2:**
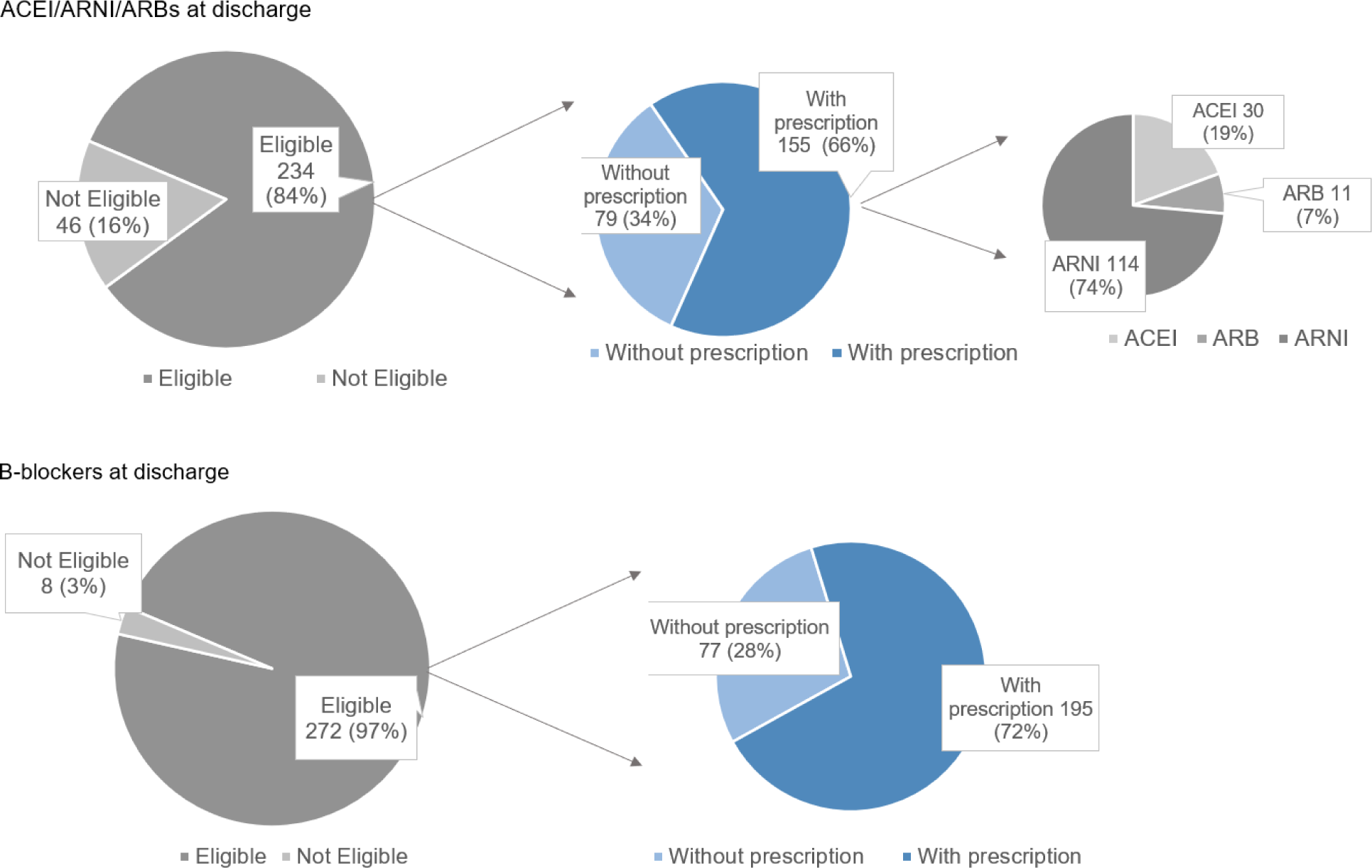
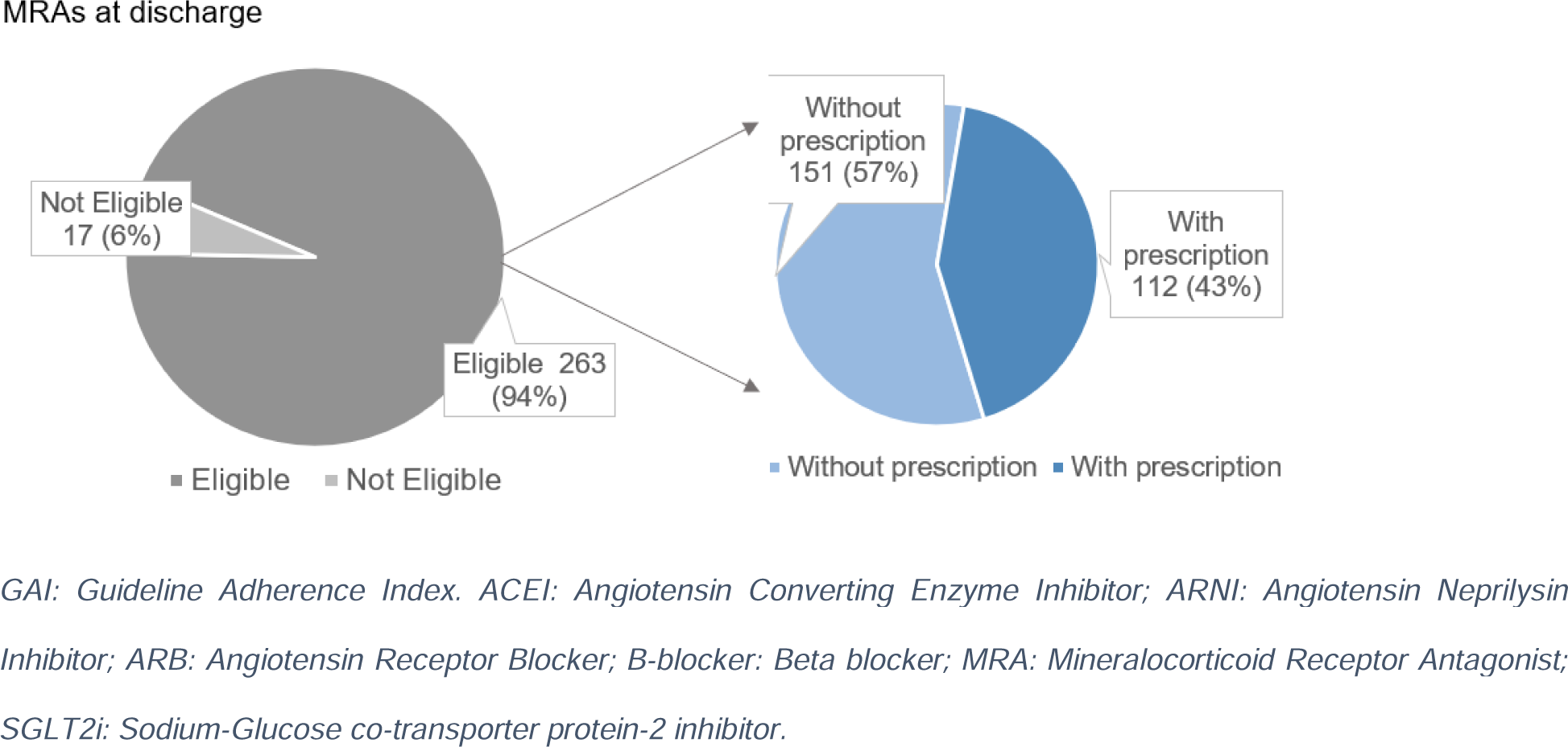
GAI at discharge.

**Figure 3:**
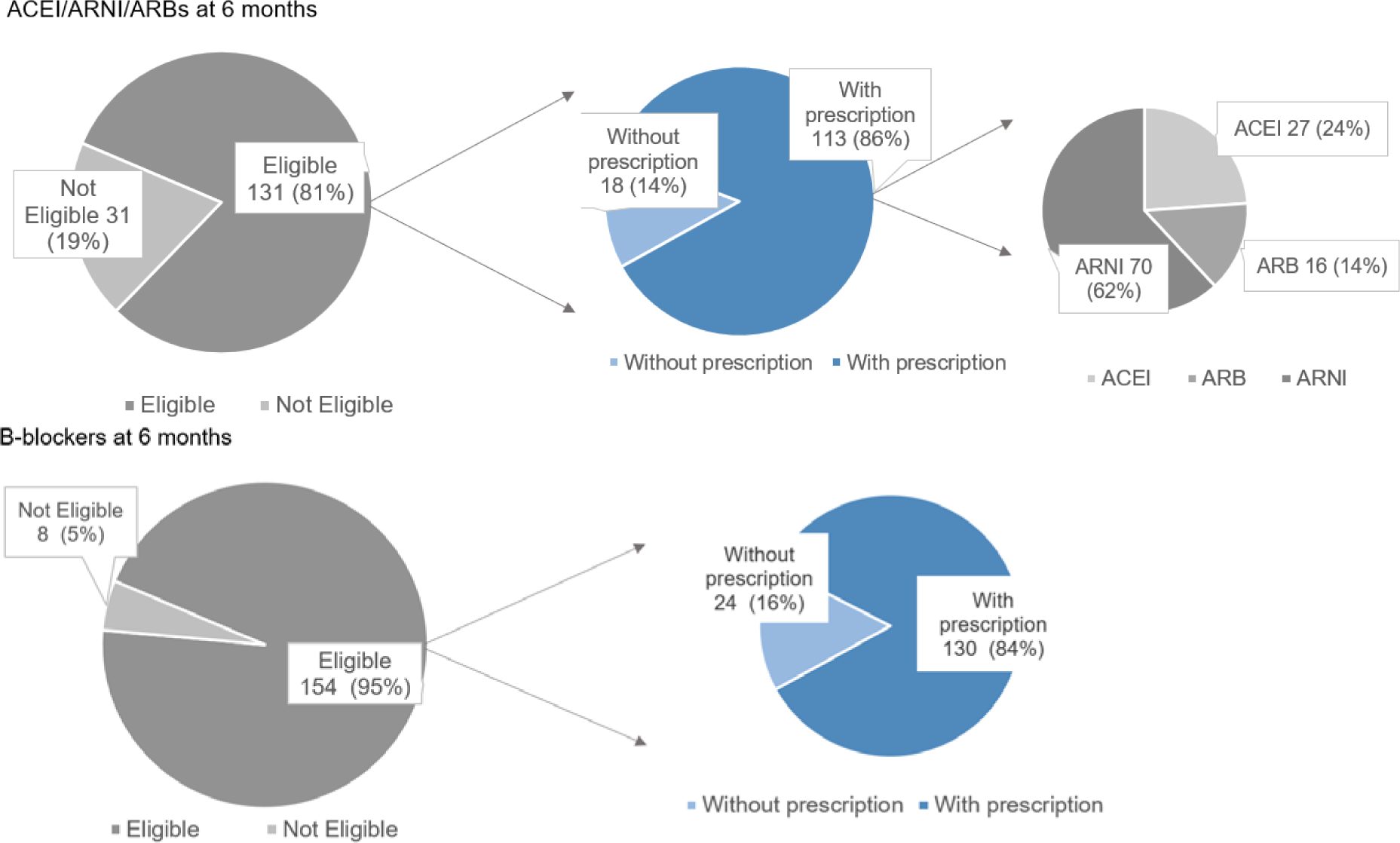
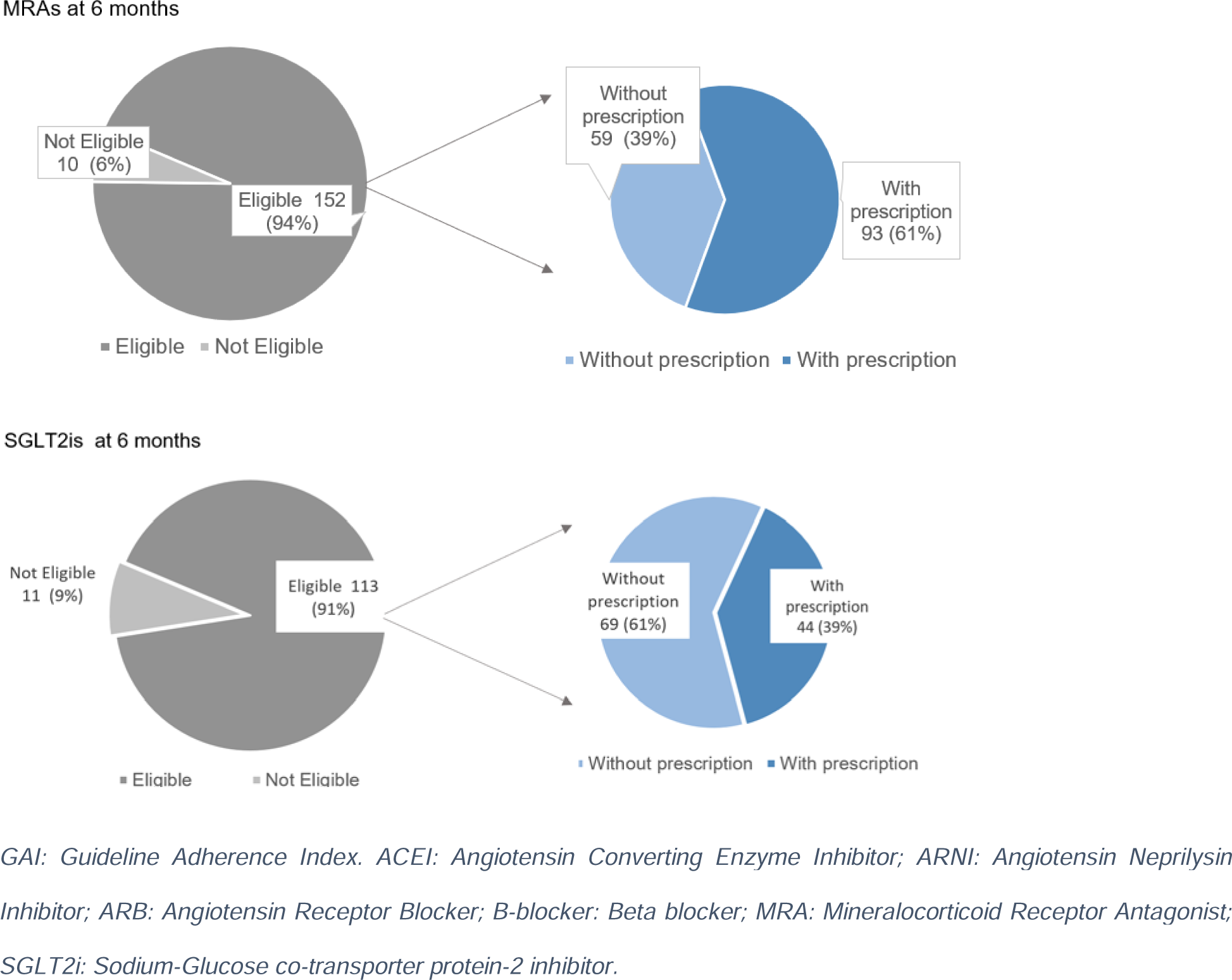
GAI at 6 months follow-up.

The overall QUALIFY scores for the 3 drug classes (i.e., ACEI/ARNI/ARBs, B-blockers, and MRAs) were 43.1% good, 33.1% moderate, and 23.8% poor. The analysis of SGLT2is prescription at 6 months showed that out of the 131 patients who were eligible, only 44 had prescriptions, resulting in a GAI of 38%, which was the lowest. Data on SGLT2is prescription at discharge was not collected because some patients had been discharged before the addition of SGLT2is to HFrEF treatment guidelines.

**Figure 4:**
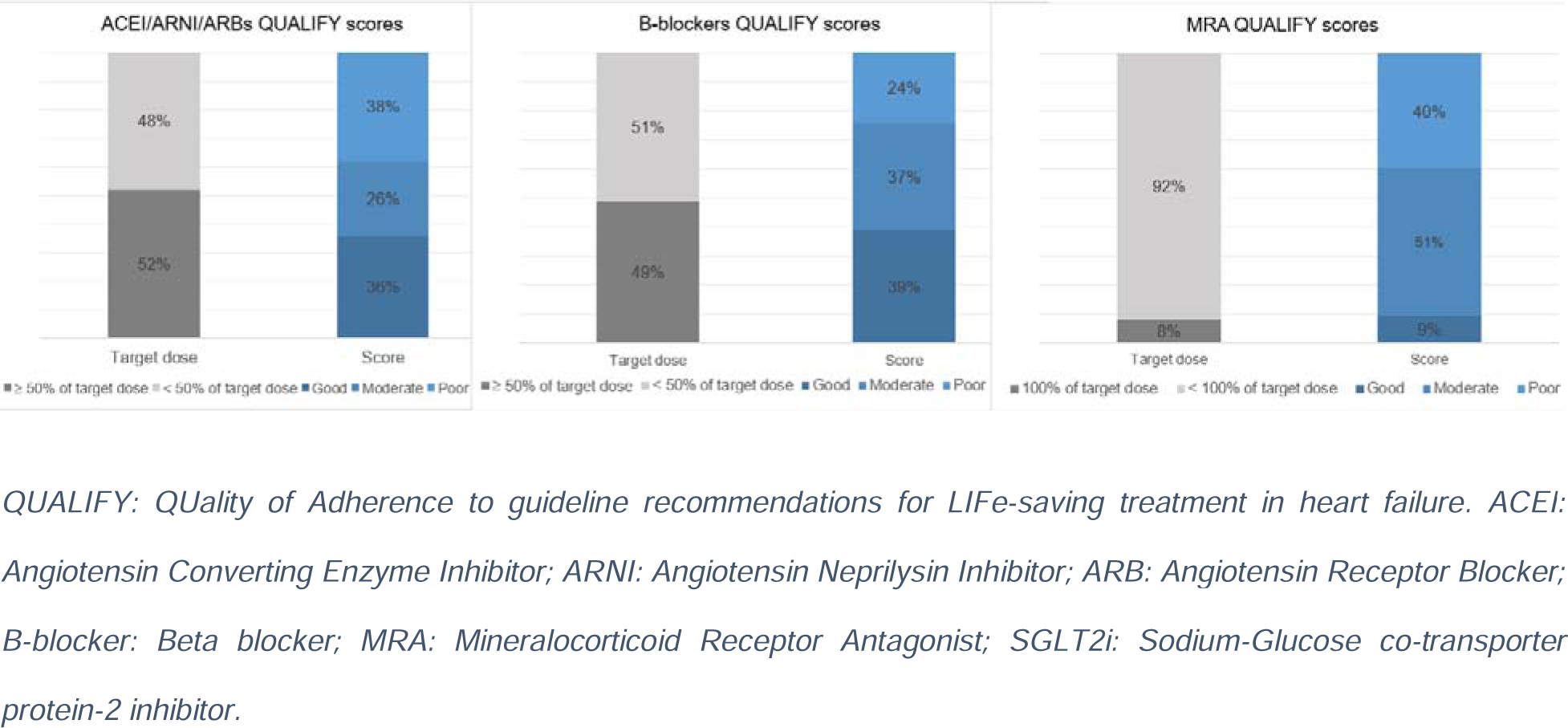
QUALIFY scores at 6 months follow-up.

### Exploratory outcomes

Multivariate analysis identified eligibility for ACEI/ARNI/ARBs at discharge as the independent factor of survival at 6 months. Eligible patients were 6 times less likely to die within 6 months compared to non-eligible patients (OR: 0.158, CI=0.041, 0.611, P=0.008). Considering the three drug classes, among eligible patients at discharge, the mortality rate was consistently lower among those with prescriptions compared with those without prescriptions at discharge, but the differences were not statistically significant for B-blockers and MRAs. Comparing eligible patients at discharge with prescriptions and those without prescriptions, the mortality rates were 2.1% vs 8.8%, 8.2% vs 9.5%, and 4.6% vs 8.1%, for ACEI/ARNI/ARBs, B-blockers, and MRAs, respectively (*Supplement V*). The analysis of independent factors associated with readmission within 6 months of follow-up found no statistically significant associations (*Supplement VI*).

## DISCUSSION

We conducted an analysis of HFrEF patients’ characteristics and physicians’ GDMT prescriptions at discharge and 6 months follow-up. Our study population has a similar mean age compared to the QUALIFY international registry (62.8 ±14.2 years vs 63.1 ±12.5 years, respectively), but younger compared to KorAHF registry (62.8 ±14.2 years vs 68.5 ±14.5 years). Our study population is predominantly black (74.2% vs <11.3% in the registries.[24], [30] Although 35% of our study population had ischemic heart disease, only 19% had documented history of MI which is much lower than 46.3% MI prevalence in QUALIFY registry.[24] This difference may signal considerable prevalence of undiagnosed MI in Kenya. In contrast to previous literature from the SSA region, IHD was the most prevalent cause of HF.[3] Patients in the current study had a prevalence of diabetes comparable to that of QUALIFY and KorAHF (36.1% vs 34.3% vs 40%, respectively), whereas the prevalence of hypertension was slightly lower (54.6% vs 64.6% vs 62.2%, respectively).

Regarding the clinical presentation, patients’ distribution according to NYHA functional classes I, II, III, and IV were similar to that of QUALIFY registry (5.2%, 56.4%, 29.9%, 8.5% vs 13%, 46%, 36%, 15% respectively). The SBP and HR were similar (125 ±23 vs 126.5 ± 20.3, however the mean HR was higher in our population (86 ±20 bpm vs 76.4 ± 14.4 bpm). The mean BMI was similar to that of QUALIFY registry (28.1 ±5.7 vs 27.9 ±5.4) but higher than that of KorAHF (28.1 ±5.7 vs 23.3 ±3.9). The LVEF was slightly lower in our study population comparable with QUALIFY and KorAHF study populations (27.6 ± 9.5% vs 31.9 ±7% vs 37.7±15.6% respectively).

Compared to published studies in SSA, the proportions of patients with prescriptions among eligible patients were worse for ACEI/ARNI/ARBs, similar or better for B-blockers, and similar for MRAs. These proportions were 66.2%, 71.8%, and 42.6%, respectively, compared to 83%, 48%, and 41%, respectively, found in a single-center study conducted in Lagos, Nigeria.[28] Another single-center study conducted in Addis Ababa, Ethiopia found these proportions to be 79.8%, 79.4%, and 49%, respectively.[29] The consistently low prescription rate of MRAs for eligible patients requires further studies on physicians’ knowledge, attitudes, and practices regarding this drug class.

The overall adherence was lower than that of QUALIFY registry. QUALIFY scores were good in 43.1%, moderate in 33.1%, and poor in 23.8% of patients, while the scores in the QUALIFY registry were good in 67%, moderate in 25% and poor 8% of patients. In the QUALIFY registry, the proportions of patients who were prescribed ≥50% of the target dose of ACEIs, ARBs, B-blockers, were 63.3%, 39.5%, and 51.8%, respectively. These proportions were lower in our study, being 51.9% for ACEI/ARNI/ARBs and 48.7% for B-blockers. Only 7.9% of patients were on 100% of MRAs target dose in our study compared to 70.4% in the QUALIFY registry.

The multivariate analysis of factors associated with mortality found that the odds of dying within 6 months were 6 times lower for patients who were eligible for ACEI/ARNI/ARBs at discharge than those who were not (OR: 0.158, CI: 0.041 - 0.611, p: 0.008). A possible explanation is that the same reasons for non-eligibility for ACEI/ARNI/ARBs are the same risk factors for death. In the current study, compared to patients eligible for ACEI/ARNI/ARBs, non-eligible patients had, lower blood pressure (SBP <90 mmHg), lower eGFR, and higher potassium levels (>5.5 mmol/L). KoAHF and other studies have reported a significant association between low eGFR and high mortality risk in HF patients.[30], [31] In the same registry, low BP (<100 mmHg) was associated with higher all-cause mortality (OR: 2.45, CI: 1.6 - 3.5), p: <0.001).

The multivariate analysis did not identify factors statistically significantly associated with readmission within 6 months. Previous studies have identified hemoglobin level, BP at admission, patient’s compliance to treatment, age, NYHA functional class, diabetes, hypothyroidism, and hypertension as independent factors for HF readmission within 6 months.[32], [33] The reason for lack of statistically significant association between readmissions and patients’ factors or HF treatment factors in our study could be low statistical power to detect these associations. A bigger sample size would potentially have more events and significant associations.

### Study limitations

The current study has several limitations. It was a retrospective study, which among other limitations, had missing data. Loss to follow-up was a significant limitation as well. The reasons for not coming for follow-up are not known and may have skewed the results at six months. Furthermore, loss to follow-up has led to limited statistical power in assessing the association between GDMT prescription and clinical outcomes at six months. However, these were secondary outcomes of the study and do not affect the primary outcome which is descriptive.

We did not capture all the possible reasons for non-prescription or non-uptitration of GDMT. We acknowledge that there are potentially more contraindications and reasons for discontinuation than those listed in the 2021 ESC guidelines. Additionally, our study does not capture physicians’ factors leading to their prescription habits. Finally, we conducted the study in a referral university hospital in the capital city, consequently, the findings are not generalizable to the whole country or region.

## CONCLUSION

Physicians’ level of adherence to prescription and up-titration of GDMT was satisfactory for ACEI/ARNI/ARBs and B-blockers, however, it was poor for SGLT2is and MRAs which had the lowest prescription rates for eligible patients. Uptitration to ≥50% of the target dose was low for all the drug classes. Exploratory analysis showed that eligibility for ACEI/ARNI/ARBs was associated with increased survival at 6 months. There is a need for regular surveys on prescription and uptitration of GDMT for HFrEF with a special attention to MRAs and SGLT2is.

## Data Availability

All data produced in the present study are available upon reasonable request to the authors.

## ACKNOWLEDGEMENTS

We would like to acknowledge the QUALIFY investigators’ contribution to the assessment of heart failure guideline adherence globally. Their work has been invaluable in advancing our understanding of the subject matter and the execution of the present study. Thank you for your hard work and dedication to this field.

