## Supplemental material for "PHYSICIANS’ ADHERENCE TO GUIDELINES ON MEDICAL THERAPY FOR HEART FAILURE AND PATIENTS’ OUTCOMES IN A KENYAN REFERRAL HOSPITAL"

Supplement I: Data extraction form

| Age (years) |  |
| --- | --- |
| Gender |  |
| Race | Black |
|  | Asian |
|  | Other |
| Reason for contact | Worsening HF |
|  | New onset HF |
|  | Planned review |
| Admission area | Critical care |
|  | Non-critical care |
| Practice | General practice |
|  | Private practice |
| Previous HF | No |
|  | <30 days |
|  | >=30 days |
| Primary cause of HF | Hypertension |
|  | Idiopathic dilated CMP |
|  | Ischemic heart disease |
|  | Other |
|  | Unknown |
| Pregnant |  |
| Past medical history | Myocardial infarction |
|  | Revascularization |
|  | No |
|  | PCI |
|  | CABG |
|  | Hypertension |
|  | Atrial fibrillation |
|  | Dilated CMP |
|  | Valve heart surgery |
|  | Major non-cardiac surgery |
|  | Rheumatic heart disease |
|  | Non-rheumatic VHD |
|  | Diabetes |
|  | COPD |
|  | Cancer |
|  | Chemotherapy |
|  | HIV |
| History of coronary angiogram | Yes |
|  | No |

| NYHA functional class | I |
| --- | --- |
|  | II |
|  | III |
|  | IV |
| BMI |  |
| BMI categories | Underweight |
|  | Normal |
|  | Overweight |
|  | Obese |
| SBP at discharge |  |
| Heart rate at discharge |  |
| LVEF |  |
| LVEF categories | ≤10% |
|  | 11 - 20% |
|  | 21 - 30% |
|  | 31 - 40% |
| ECG | Sinus rhythm |
|  | Atrial fibrillation |
|  | Paced rhythm |
|  | Other |
| Chest x-ray | Not done |
|  | Normal |
|  | Pulmonary edema |
|  | Cardiomegaly |
|  | Pulmonary edema and cardiomegaly |
| Covid status | Positive |
|  | Negative |
|  | Unknown |
| eGFR |  |
| Potassium |  |
| Cardiac rehabilitation | No |
|  | Initiated only |
|  | Completed |

| **6 months follow-up** |  |
| --- | --- |
| SBP at 6 months |  |
| Heart rate at 6 months |  |
| NYHA functional class at 6 months | I |
|  | II |
|  | III |
|  | IV |
| eGFR at 6 months |  |
| Serum potassium at 6 months |  |
| Cardiac rehabilitation at 6 months | No |
|  | Initiated only |
|  | Completed |
| Readmission within 6 months | No |
|  | Once |
|  | Twice |
|  | Thrice |
| Mortality within 6 months | Died |
|  | Alive |
| Cause of death | CV |
|  | Non-CV |
|  | Unknown |

| **Medications** |  |  |  |  |
| --- | --- | --- | --- | --- |
|  |  | **Name** | **Dose (Index discharge)** | **Dose (month 6)** |
| ACEIs or ARBs | Medications | Captopril |  |  |
|  |  | Enalapril |  |  |
|  |  | Lisinopril |  |  |
|  |  | Ramipril |  |  |
|  |  | Trandolapril |  |  |
|  |  | Candesartan |  |  |
|  |  | Losartan |  |  |
|  |  | Valsartan |  |  |
|  | Contraindications / Reason for discontinuation | Pregnancy |  |  |
|  |  | Known b/l renal artery stenosis |  |  |
|  |  | Known allergic reaction |  |  |
|  |  | Angioedema |  |  |
|  |  | K+ rise >5.5 mmol/L, 100% creatinine rise or >310 umol/L |  |  |
| ARNI | Medication | Sacubitril/valsartan |  |  |
|  | Contraindications /  Reason for discontinuation | Pregnancy / breastfeeding |  |  |
|  |  | eGFR < 30 ml/min/1.73 m2 |  |  |
|  |  | Symptoms of hypotension or SBP <90 mmgHg |  |  |
|  |  | History of angioedema |  |  |
|  |  | Known allergic reaction |  |  |
|  |  | K+ rise >5.5 mmol/L, or eGFR drop to <30 ml/min/1.73m2 |  |  |
| Beta-blockers | Medication | Bisoprolol |  |  |
|  |  | Carvedilol |  |  |
|  |  | Metoprolol |  |  |
|  |  | Nebivolol |  |  |
|  | Contraindications /  Reasons for discontinuation | 2nd or 3rd degree AV block (in the absence of permanent pacemaker |  |  |
|  |  | Critical limb ischemia |  |  |
|  |  | Asthma (relative contraindication) |  |  |
|  |  | Known allergic reaction |  |  |
|  |  | Bradycardia <50 bpm |  |  |
|  |  | Persisting signs of congestion, hypotension (<90 mmHg) |  |  |
| MRAs | Medication | Eplerenone |  |  |
|  |  | Spironolactone |  |  |
|  | Contraindications /  Reason for discontinuation | Known allergic reaction |  |  |
|  |  | K+ rise >6 mmol/L, or creatinine to 310 umol/l, eGFR drop to <20 ml/min/1.73m2 |  |  |

| SGLT2is | Medication | Dapagliflozin |
| --- | --- | --- |
|  |  | Empagliflozin |
|  | Contraindications /  Reason for discontinuation | Known allergic reaction |
|  |  | Pregnancy/risk of pregnancy and breastfeeding period |
|  |  | Symptoms of hypotension or SBP <95mmHg |
|  |  | eGFR <20 ml/min/1.73m2 |
| **Outcomes** |  | |
| Number of readmissions |  | |
| Death | Yes/No | |
| Cause of death | CV / Non-CV | |

Supplement II: GDMT target doses

| Medications | Name | Target dose |
| --- | --- | --- |
| ACEIs | Captopril | 50 mg tid |
|  | Enalapril | 20 mg bd |
|  | Lisinopril | 35 mg bd |
|  | Ramipril | 10 mg od |
|  | Trandolapril | 4 mg od |
|  | Perindopril | 16 mg od |
| ARNI | Sacubitril/Valsartan | 200 mg bd |
| ARBs | Candesartan | 32 mg od |
|  | Valsartan | 160 mb bd |
|  | Losartan | 100 mg od |
| B-blockers | Bisoprolol | 10 mg od |
|  | Carvedilol | 25 mg bd (50 bd if > 85kgs) |
|  | Metoprolol | 200 mg od |
|  | Nebivolol | 10 mg od |
| MRAs | Eplerenone | 50 mg od |
|  | Spironolactone | 50 mg od |
| SGLT2is | Dapagliflozin | 10 mg od |
|  | Empagliflozin | 10 mg od |

Supplement III: GAI and QUALIFY score calculation

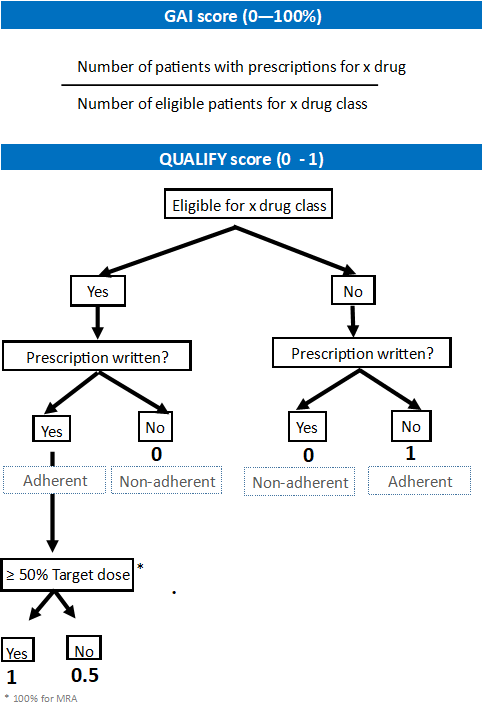

Supplement IV: Detailed GAI and QUALIFY scores at discharge and at 6 months

|  | **At discharge** | | **At 6 months** | |
| --- | --- | --- | --- | --- |
| ACEI/ARNI/ARBs prescribed | ACEIs: | 30 (10.7) | 27 (16.7) | |
|  | ARBs: | 11 (3.9) | 16 (9.9) | |
|  | ARNI: | 114 (40.7) | 70 (43.2) | |
|  | Total: | 155 (55.4) | 113 (69.7) | |
| Eligible for ACEI/ARNI/ARBs | 234 (83.6) | | 131 (80.8) | |
| GAI for ACEI/ARNI/ARBs | 155/234 (66.2) | | 113/131 (86.3) | |
| Prescribed ACEI/ARNI/ARBs (n=104) |  | | < 50% of target dose: | 50 (48.1) |
|  |  | | ≥ 50% of target dose: | 54 (51.9) |
| QUALIFY for ACEI/ARNI/ARBs (n=148) |  | | Good: | 53 (35.8) |
|  |  | | Moderate: | 39 (26.6) |
|  |  | | Poor: | 56 (37.8) |
| B-blockers prescribed | 195 (69.6) | | 130 (80.2) | |
| Eligible for B-blockers | 272 (97.1) | | 154 (95.1) | |
| GAI for B-blockers | 195/272 (71.8) | | 130/154 (84.4) | |
| Prescribed B-blocker dose (n=117) |  | | < 50% of target dose: | 60 (51.3) |
|  |  | | ≥ 50% of target dose: | 57 (48.7) |
| QUALIFY for B-blockers (n=148) |  | | Good: | 57 (38.5) |
|  |  | | Moderate: | 55 (37.2) |
|  |  | | Poor: | 36 (24.3) |
| MRAs prescribed | 112 (40.6) | | 93 (57.4) | |
| Eligible for MRAs | 263 (93.9) | | 152 (93.8) | |
| GAI for MRAs | 112/263 (42.6) | | 93/152 (61.2) | |
| Prescribed MRA dose at 6 months (n=89) |  | | < 100% of target dose: | 82 (92.1) |
|  |  | | 100% of target dose: | 7 (7.9) |
| QUALIFY for MRAs (n=158) |  | | Good: | 15 (9.5) |
|  |  | | Moderate: | 80 (50.6) |
|  |  | | Poor; | 63 (39.9) |
| SGLT2is prescribed at 6 months (n=124) |  | | 44 (35.5) | |
| Eligible for SGLT2is at 6 months (n=124) |  | | 113 (91.1) | |
| GAI for SGLT2is at 6 months (n=124) |  | | 44/113 (38) | |

Supplement V: Factors associated with mortality within 6 months of follow-up

| **Univariate binary logistic regression of factors associated with mortality within 6 months** | | | | |
| --- | --- | --- | --- | --- |
|  |  | OR | 95% CI | p Value |
| Age (years) |  | 0.995 | (0.960, 1.032) | 0.783 |
| Gender | Male | 0.421 | (0.146, 1.212) | 0.109 |
| Race | Black | 1.4 | (0.422, 4.646) | 0.582 |
|  | Asian | 0 | (0,) | 0.999 |
| Reason for contact | Worsening HF | 0.507 | (0.51, 5.049) | 0.562 |
|  | New onset HF | 1.547 | (0.178, 13.45) | 0.693 |
|  | Planned review | 0 | (0,) | 0.998 |
| Admission area | Critical care | 1.233 | (0.426, 3.571) | 0.699 |
| Practice | University Hospital | 0.636 | (0.208, 1.946) | 0.428 |
| Previous HF | < 30 days | 0.58 | (0.143, 2.344) | 0.444 |
|  | >= 30 days | 0.394 | (0.122, 1.269) | 0.119 |
| Cause of HF | Idiopathic DCM | 0.933 | (0.230,3.794) | 0.923 |
|  | Ischemic HD | 0.25 | (0.046, 1.359) | 0.109 |
|  | Other | 0.897 | (0.240, 3.363) | 0.872 |
|  | Unknown | 0 | (0,) | 0.999 |
| MI |  | 0.638 | (0.138, 2.927) | 0.566 |
| HTN |  | 0.648 | (0.230, 1.823) | 0.648 |
| A-fib |  | 1.782 | (0.533, 5.956) | 0.348 |
| DM | Type 1 DM | 1.063 | (0.215,5.247) | 0.94 |
|  | T2DM - on insulin | 4.074 | (0.934,17.768) | 0.062 |
|  | T2DM on oral treatment | 1.111 | (0.225, 5.498) | 0.897 |
|  | T2DM on diet | 0 | (0,) | 0.999 |
| COPD |  | 4.571 | (0.812, 25.745) | 0.085 |
| Non-rheumatic VHD |  | 4.571 | (0.812, 25.745) | 0.085 |
| DCM |  | 0.773 | (0.641, 7.32) | 0.213 |
| Cancer |  | 1.308 | (0.153, 11.179) | 0.806 |
| Chemotherapy |  | 0 | (0,) | 0.999 |
| HIV |  | 0 | (0,) | 0.999 |
| Heart valve surgery |  | 1.767 | (0.199, 15.663) | 0.609 |
| Revascularization | CABG | 2.148 | (0.415, 11,112) | 0.362 |
|  | PCI | 1.742 | (0.549, 5.524) | 0.346 |
| Major non-cardiac surgery |  | 0.512 | (0.064, 4.1) | 0.528 |
| NYHA | I | 0 | (0,) | 0.999 |
|  | II | 0.903 | (0.177, 4.603) | 0.902 |
|  | III | 0.464 | (0.070, 3.086) | 0.464 |
| BMI | I | 0 | (0,) | 0.999 |
|  | II | 2.419 | (0.571, 10.245) | 0.23 |
|  | III | 1.421 | (0.324, 6.231) | 0.642 |
| SBP |  | 1.005 | (0.985, 1.025) | 0.649 |
| PR |  | 1.006 | (0.984, 1.029) | 0.585 |
| LVEF |  | 1.005 | (0.953, 1.060) | 0.85 |
| eGFR |  | 0.987 | (0.970, 1.005) | 0.149 |
| Potassium |  | 1.928 | (0.964, 3.854) | 0.063 |
| ECG | Atrial fibrillation | 1.133 | (0.291, 4.408) | 0.857 |
|  | Other non-sinus rhythm | 0.887 | (0.182, 4.324) | 0.882 |
|  | Paced | 1.133 | (0.130, 9.883) | 0.91 |
| Cardiac rehab initiated |  | 0.844 | (0.093, 7.672) | 0.88 |
| Eligible for ACEI/ARNI/ARB |  | 0.113 | (0.038, 0.337) | <0.001 |
| ACEI/ARNI/ARB prescribed |  | 0.48 | (0.170, 1.354) | 0.166 |
| Eligible for beta blocker |  | 0.373 | (0.039, 3.551) | 0.391 |
| Beta blocker prescribed |  | 0.929 | (0.283, 3.044) | 0.903 |
| Eligible for MRA |  | 0.112 | (0.031, 0.4) | 0.001 |
| MRA prescribed |  | 0.359 | (0.098, 1.31) | 0.121 |
| **Multivariate binary logistic regression of independent factors associated with mortality within 6 months** | | | | |
| Eligible for ACEI/ARNI/ARB |  | 0.158 | (0.041, 0.611) | 0.008 |
| Eligible for MRA |  | 0.492 | (0.101, 2.396) | 0.38 |
| **Crosstabulation: GDMT prescription rate at discharge and mortality at 6 months** | | | |  |
|  |  | Alive (%) | Dead (%) |  |
| ACEI/ARNI/ARB prescribed for eligible patient | Yes | 92 (97.9) | 2 (2.1) |  |
|  | No | 52 (91.2) | 5 (8.8) |  |
| Beta blocker prescribed for eligible patient | Yes | 123 (91.8) | 11 (8.2) |  |
|  | No | 38 (90.5) | 4 (9.5) |  |
| MRA prescribed for eligible patient | Yes | 62 (95.4) | 3 (4.6) |  |
|  | No | 91 (91.9) | 8 (8.1) |  |

Supplement VI: Factors associated with readmission within 6 months of follow-up.

| **Univariate binary logistic regression of factors associated with readmission within 6 months** | | | | |
| --- | --- | --- | --- | --- |
|  |  | OR | 95% CI | P Value |
| Age (years) |  | 0.995 | (0.960, 1.032) | 0.783 |
| Gender | Male | 0.421 | (0.146, 1.212) | 0.109 |
| Race | Black | 0.434 | (0.099, 1.905) | 0.269 |
|  | Asian | 0.508 | (0.098, 2.620) | 0.418 |
| Reason for contact | Worsening HF | 2.25 | (0.421, 12.028) | 0.343 |
|  | New onset HF | 3.22 | (0.598, 17.358) | 0.173 |
|  | Planned review | 1.25 | (0.185, 8.444) | 0.819 |
| Admission area | Critical care | 1.333 | (0.666, 2.669) | 0.417 |
| Practice | University Hospital | 1.21 | (0.536, 2.753) | 0.642 |
| Previous HF |  |  |  | 0.084 |
|  | < 30 days | 0.583 | (0.228, 1.495) | 0.264 |
|  | > 30 days | 0.424 | (0.199, 0.904) | 0.026 |
| Cause of HF |  |  |  | 0.191 |
|  | Idiopathic DCM | 2.369 | (0.819, 6.855) | 0.112 |
|  | Ischemic HD | 1.29 | (0.495, 3.360) | 0.603 |
|  | Other | 2.933 | (1.075, 8.001) | 0.036 |
|  | Unknown | 2.2 | (0.270, 17.924) | 0.461 |
| MI |  | 1.149 | (0.489, 2.697) | 0.75 |
| HTN |  | 0.88 | (0.455, 1.704) | 0.705 |
| A-fib |  | 0.798 | (0.321, 1.983) | 0.627 |
| DM |  |  |  | 0.32 |
|  | Type 1 DM | 1.19 | (0.431, 3.281) | 0.737 |
|  | T2DM - on insulin | 10.41 | (1.232, 87.973) | 0.031 |
|  | T2DM on oral treatment | 1.217 | (0.461, 3.21) | 0.692 |
|  | T2DM on diet | 0 | (0,) | 0.999 |
| COPD |  | >999 | (0,) | 0.999 |
| Non-rheumatic VHD |  | >999 | (0,) | 0.999 |
| DCM |  | 1.882 | (0.738, 4.798) | 0.185 |
| Cancer |  | 0.171 | (0.020, 1.424) | 0.102 |
| Chemotherapy |  | 0.245 | (0.028, 2.154) | 0.205 |
| HIV |  | 1.29 | (0.079, 21.041) | 0.858 |
| Heart valve surgery |  | 0.852 | (0.138, 5.263) | 0.854 |
| Revascularization | CABG | 1.341 | (0.366, 4.918) | 0.658 |
|  | PCI | 1.104 | (0.492, 2.477) | 0.81 |
| Major non-cardiac surgery |  | 0.189 | (0.041, 0.876) | 0.033 |
| NYHA | I | 0.457 | (0.076, 2.764) | 0.394 |
|  | II | 0.337 | (0.089, 1.277) | 0.11 |
|  | III | 0.639 | (0.159, 2.569) | 0.528 |
| BMI |  |  |  | 0.177 |
|  | I | 0.955 | (0.123, 7.408) | 0.965 |
|  | II | 0.955 | (0.394, 2.314) | 0.918 |
|  | III | 0.441 | (0.194, 0.998) | 0.05 |
| SBP |  | 1.002 | (0.989, 1.016) | 0.743 |
| PR |  | 1.008 | (0.993, 1.024) | 0.288 |
| LVEF |  | 1.006 | (0.972, 1.041) | 0.729 |
| eGFR |  | 0.995 | (0.985, 1.005) | 0.364 |
| Potassium |  | 0.842 | (0.479, 1.483) | 0.553 |
| ECG | Atrial fibrillation | 0.622 | (0.240, 1.612) | 0.329 |
|  | Other non-sinus rhythm | 0.742 | (0.264, 2.087) | 0.572 |
|  | Paced | 1.167 | (0.275, 4.953) | 0.834 |
| Cardiac rehab initiated |  | 0.64 | (0.113, 3.619) | 0.614 |
| Eligible for ACEI/ARNI/ARB |  | 1.127 | (0.403, 3.148) | 0.82 |
| ACEI/ARNI/ARB prescribed |  | 0.956 | (0.484, 1.886) | 0.896 |
| Eligible for beta blocker |  | 0.381 | (0.034, 4.302) | 0.435 |
| Beta blocker prescribed |  | 1.346 | (0.613, 2.957) | 0.459 |
| Eligible for MRA |  | 0.769 | (0.15, 3.947) | 0.753 |
| MRA prescribed |  | 0.516 | (0.256, 1.038) | 0.063 |
| **Multivariate binary logistic regression of independent factors associated with readmission within 6 months** | | | | |
|  |  | OR | 95% C.I. | p value |
| Previous HF |  |  |  | 0.192 |
|  | < 30 days | 0.632 | (0.199, 2.005) | 0.436 |
|  | >= 30 days | 0.41 | (0.154, 1.089) | 0.074 |
| Cause of heart failure |  |  |  | 0.187 |
|  | Idiopathic DCM | 3.582 | (0.936, 13.701) | 0.062 |
|  | Ischemic HD | 1.774 | (0.524, 6.003) | 0.357 |
|  | Other | 3.813 | (1.17, 12.426) | 0.026 |
|  | Unknown | 4.924 | (0.313, 77.369) | 0.257 |
| History of Diabetes |  |  |  | 0.276 |
|  | Type 1 DM | 1.938 | (0.559, 6.726) | 0.297 |
|  | T2DM - on insulin | 12.762 | (1.198, 135.963) | 0.035 |
|  | T2DM on oral treatment | 1.537 | (0.491, 4.81) | 0.46 |
|  | T2DM on diet | 0 | (0,) | 0.999 |
| History of major non-card surg(1) |  | 0.369 | (0.071, 1.913) | 0.235 |
| BMI categories |  |  |  | 0.455 |
|  | I | 0.558 | (0.046, 6.736) | 0.646 |
|  | II | 0.902 | (0.321, 2.535) | 0.845 |
|  | III | 0.5 | (0.201, 1.244) | 0.136 |
